# Cluster analysis of angiotensin biomarkers to identify antihypertensive drug treatment in population studies

**DOI:** 10.1101/2022.06.21.22276541

**Authors:** Arisido M Woldeyes, Foco Luisa, Shoemaker Robin, Melotti Roberto, Delles Christian, Gögele Martin, Barolo Stefano, Baron Stephanie, Azizi Michel, Dominiczak Anna, Zennaro M Christina, Pramstaller P Peter, Poglitsch Marko, Pattaro Cristian

## Abstract

**Background:** Hypertension is a leading cause of death worldwide. Population-based studies offer an opportunity to assess the effectiveness of anti-hypertensive drugs (AHD) in real-world scenarios. However, lack of quality AHD documentation, especially when electronic health record linkage is unavailable, leads to reporting and classification bias. Here we assessed to which extent Renin-Angiotensin-Aldosterone System (RAAS) biomarkers can identify AHD treatments in the general population.

**Method:** Angiotensin I, angiotensin II and aldosterone levels were simultaneously determined through mass-spectrometry analysis in 800 participants of the Cooperative Health Research In South Tyrol (CHRIS) study with documented AHD treatment. We conducted unsupervised cluster analysis, assessing agreement, sensitivity and specificity of the resulting clusters against known AHD treatment. Through lasso penalized regression we identified clinical characteristics associated with RAAS biomarkers, accounting for the effects of cluster and treatment classifications.

**Results:** We identified three well-separated clusters: cluster 1 (n=444) preferentially including individuals not receiving RAAS-targeting AHD; cluster 2 (n=235) identifying angiotensin type 1 receptor blockers (ARB) users (weighted kappa κ_w_=74%; sensitivity=73%; specificity=83%); and cluster 3 (n=121) well discriminating angiotensin-converting enzyme inhibitors (ACEi) users (κ_w_=81%; sensitivity=55%; specificity=90%). Individuals in clusters 2 and 3 had higher frequency of diabetes as well as higher fasting glucose and BMI levels. Age, sex and kidney function were strong predictors of the RAAS biomarkers independently of the cluster structure.

**Conclusions:** Unsupervised clustering of angiotensin I, angiotensin II and aldosterone is a viable technique to identify individuals on ACEi and ARB AHD treatment outside of a controlled clinical setting.

## Introduction

Hypertension is a leading cause of death worldwide [1] and a primary risk factor for various comorbidities [2]. Despite widespread availability of anti-hypertensive drugs (AHD), the global prevalence of uncontrolled hypertension in treated individuals remains high, surpassing 50% in low-income countries [3]. Drugs that suppress the renin-angiotensin-aldosterone system (RAAS) include angiotensin-converting enzyme inhibitors (ACEi) and angiotensin receptor blockers (ARB), either monotherapy or combined with a diuretic. Combinations of an ACEi or an ARB with a calcium channel blocker or a diuretic is the recommended first-line treatment for hypertension [4].

Population-based epidemiological studies provide an opportunity to assess AHD effectiveness in non-clinical contexts. While these studies generally have a much larger scale than clinical studies, they often lack high-quality documentation of the AHD treatment, especially when linkage to electronic health records is not available. In the absence of efficient drug information retrieval systems, treatment self-reporting is subject to recall and classification bias [5,6]. On the other hand, sample biobanking guarantees the possibility to measure extensive sets of molecular biomarkers afterwards and, for example, to reconstruct the most likely AHD treatment *a posteriori*. Recent advances in drug and metabolite screening via mass spectrometry was shown to improve accuracy of the assessment of underlying treatment [7], and novel methods have been validated to simultaneously measure angiotensin I, angiotensin II and aldosterone in biobanked blood samples [8].

In the present study, we quantified angiotensin I, angiotensin II and aldosterone using the RAAS-Triple A assay [8] in biobanked serum samples from a subset of 800 participants from the Cooperative Health Research In South Tyrol (CHRIS) study [9], where AHD classification was constructed accurately through automatic drug package barcode scanning upon participation.

We investigated whether unsupervised cluster analysis of the three biomarkers may help identify the undertaken AHD treatment. We evaluated the agreement between estimated clusters and the objective classification obtained via drug box barcode scanning. The resulting clusters were characterized based on available clinical information. Finally, to identify possible reasons of imperfect classification, we assessed which clinical characteristics, among those typically associated with different AHD treatments, were related to angiotensin I, angiotensin II and aldosterone while accounting for the effects of the clustering and the AHD treatments.

## Methods

### Study design and participants

This study was based on the CHRIS study, a single-center, population-based study designed to investigate the molecular, behavioral and environmental determinants of human health, whose baseline assessment was carried out between 2011 and 2018 [9,10]. Blood was collected from CHRIS study participants following overnight fasting. After immediate pre-analytical sample processing, sample storage at –80°C was performed as described in [9,10]. Health-related information was collected through either self- or interviewer-administered interviews based on standardized electronic questionnaires. Participants were requested to bring any boxes of medications taken in the preceding week to the study center. Drug information was retrieved via scanning the drug box barcodes, automatically classified according to the Farmadati database (https://www.farmadati.it/), and stored in the CHRIS database.

At the time when the present study was set up, the CHRIS study included N=6075 participants. Budget limitations allowed to measure the RAAS biomarkers on a random sample of 800 samples. Taking into account that the smallest treatment group included 100 individuals, we sampled 8 age- and sex-matched groups based on the AHD treatment: 1) normotensive; 2) untreated hypertensive; 3) participants taking other drugs not prescribed as AHD (referred to as *non-AHD*); 4) participants on ACEi monotherapy; 5) participants on ACEi combined with diuretics (*ACEi+diuretics*); 6) participants on ARB monotherapy; 7) participants on ARB in combination with diuretics (*ARB+diuretics*); and 8) participants on beta blocker monotherapy treatment (*Beta blockers*).

### Clinical characteristics

Blood pressure (BP) was measured in supine position with the Omron digital automatic BP Monitor M10-IT at the end of a 20-minute resting electrocardiogram. The mean of three measurements taken at 2 minutes intervals was recorded. Hypertension was defined as: reporting the use of an AHD (ATC codes starting with C02, C03, C04, C07, C08, and C09) or having a diastolic BP (DBP) of ≥90 mmHg or a systolic BP (SBP) of ≥140 mmHg, according to established guidelines [4]. Diabetes mellitus (DM) was defined as a positive answer to the question “*Do you have diabetes mellitus?*” or the reporting of glucose-lowering drugs (ATC codes: A10) or by measured levels of glycated haemoglobin (HbA1c) ≥6.5% (7.8 mmol/L) or glucose ≥126 mg/dl (7 mmol/L) [11]. Estimated glomerular filtration rate (eGFR) was obtained from serum creatinine using the CKD-EPI formula [12]. Serum levels of total cholesterol, cortisol, potassium and sodium were determined as previously described [10].

### Quantification of RAAS biomarkers

Equilibrium Angiotensin I, angiotensin II and aldosterone levels were simultaneously determined using RAAS Triple A testing (Attoquant Diagnostics GmbH, Vienna, Austria) via liquid chromatography combined with tandem mass spectrometry (LC-MS/MS) analysis as previously described [8]. Briefly, equilibration of serum samples was performed at 37 °C for one hour, followed by stabilization through addition of an enzyme inhibitor cocktail. Samples were spiked with stable isotope-labeled internal standards for each analyte, and subjected to C-18-based solid-phase-extraction followed by LC-MS/MS analysis using a reversed-phase analytical column operating in line with a Xevo TQ-S triple quadruple mass spectrometer (Waters). Internal standards were used to correct for peptide recovery of the sample preparation procedure for each analyte in each individual sample.

The biomarkers were quantified from integrated chromatograms considering the corresponding response factors determined in appropriate calibration curves in serum matrix, on condition that integrated signals exceeded a signal-to-noise ratio of 10. The lower limits of quantification for angiotensin I, angiotensin II and aldosterone, defined as the lowest concentrations tested showing a coefficient of variation (CV) < 20% according to FDA criteria, are 5 pg/ml for each of the three biomarkers, corresponding to 3.9, 4.8 and 13.9 pmol/L, respectively. At 50 pmol/L, the inter-assay CVs for the three biomarkers are 10.2%, 6.1%, and 7.9%, respectively, while the corresponding intra-assay CVs are 8.6%, 4.4%, and 5.2%, respectively.

### Statistical analyses

The distributions of angiotensin I, angiotensin II and aldosterone were skewed to the left, hence they were log-transformed to achieve normality (Supplementary Figure 1).

First, to determine whether the three RAAS biomarkers could identify participants in different AHD groups, we conducted K-means unsupervised cluster analysis [13,14] by assigning each observation to one of k groups based on a similarity feature computed from the biomarkers’ covariance matrix. Cluster membership is computed as the sum of the squared distance between data points and the cluster center using the Euclidean distance [15,16]. Principal components (PCs) were then used to inspect the identified clusters. The k-means method was chosen after comparison with the alternative unsupervised machine learning approaches such as hierarchical and fuzzy clustering [17]. Selection of the best method as well as the optimal number of clusters was based on the *Silhouette* score [18], which was evaluated for a number of clusters between 2 and 6.

We assessed the agreement between AHD treatment identified by the clusters and the eight groups using the weighted kappa (κ_w_) inter-agreement coefficient [19]. κ_w_ is a modification of the Cohen’s index [20] to deal with chance agreement between classifiers, and defined based on conditional probability that two classifiers will agree given that disagreement will occur by chance. Computational details are provided in [19]. Sensitivity and specificity were estimated considering the objective AHD classification obtained by the drug box barcode scanning as the gold standard.

Next, we assessed differences of the clinical characteristics among the clusters using one□way analysis of variance (ANOVA) or chi□squared test where appropriate.

Finally, we fitted a lasso penalized regression model [21] to assess whether any clinical predictor could explain the residual variance of each RAAS biomarker, which was not explained by the clusters or the treatment. We fitted a model for each biomarker as the response variable, setting clinical characteristics as fixed-effect predictors and the identified clusters and the treatment group as random-effect terms. The rationale to introducing the identified clusters and treatment groups as random effect was to capture the residual variability not accounted by the fixed effect predictors.

The lasso penalization was applied to obtain a parsimonious predictive model that should not suffer from the between-predictor pairwise correlation (Supplementary Figure 2): coefficients are constrained by imposing a penalty to drop the less influential predictors from the model by shrinking their coefficients to zero [21,22]. The penalty level was tuned by selecting a penalty parameter *λ* using k-fold cross-validation (CV), with the aim to minimize the mean squared error (MSE). We set k=8 and the smallest MSE was observed at *λ*=0.03, 0.02 and 0.01, for angiotensin I, angiotensin II and aldosterone, respectively (Supplementary Figure 3).

The statistical significance level was set at 0.05 in all analyses. All analyses were performed with the R software v4.0.5 (https://www.r-project.org), using the packages *stats* v4.2.0 and *cluster* v2.1.2 (https://CRAN.R-project.org/package=cluster) [23] for cluster analysis and *glmnet* v4.1.3 and *glmmLasso* v1.5.1 (https://CRAN.R-project.org/package=glmmLasso) [24] for penalized regression analysis.

## Results

Clinical characteristics of each group are described in Table 1. Study participants were 43 to 90 years old, and 54% were females. Normotensive participants had the lowest SBP and DBP levels, while the hypertensive group had the highest BP levels. In the treatment groups, mean BP was within the control limits, with exception of ACEi users, whose mean SBP levels were >140 mmHg. AHD treatment groups showed larger BMI levels compared to normotensive individuals, with maximal BMI levels in the ARB+diuretics group. Additional clinical characteristics are reported in Supplementary Table 1. Figure 1 illustrates the distributions of the three RAAS biomarkers across AHD groups. Angiotensin I, angiotensin II and aldosterone showed a similar joint profile in all AHD groups that do not include RAAS-targeting treatment (normotensive, hypertensive, non-AHD, and Beta blockers). In contrast, more elevated angiotensin I and depleted aldosterone levels were observed in the ACEi, ACEi+diuretics, ARB, and ARB+diuretics groups. Angiotensin II was depleted in the ACEi groups, and elevated in the ARB groups.

**Table 1:** Characteristics of the 800 participants by AHD treatment group.

| AHD group | N | Sex<br>M/F | Age, years<br>mean(SD) | SBP, mmHg<br>mean(SD) | DBP, mmHg<br>mean(SD) | BMI, kg/m<sup>2</sup><br>mean(SD) |
| --- | --- | --- | --- | --- | --- | --- |
| Normotensive | 101 | 46/55 | 64.9(7.2) | 122.3(9.4) | 78.7(6.4) | 25.9(3.4) |
| Hypertensive | 100 | 46/54 | 62.1(7.8) | 146.7(12.2) | 93.0(7.2) | 27.2(4.4) |
| Non-AHD | 100 | 46/54 | 68.8(10.1) | 131.2(18.4) | 81.0(8.3) | 26.3(4.3) |
| Beta blockers | 100 | 46/54 | 66.2(9.0) | 134.6(16.8) | 83.7(8.9) | 28.2(4.8) |
| ACEi | 99 | 45/54 | 68.7(10.0) | 142.5(18.0) | 85.5(9.4) | 28.7(4.2) |
| ACEi+diuretics | 100 | 47/53 | 68.6(10.0) | 138.1(18.7) | 84.0(8.2) | 29.8(4.3) |
| ARB | 98 | 45/53 | 65.2(9.5) | 134.9(15.0) | 84.7(8.4) | 28.6(4.8) |
| ARB+diuretics | 102 | 47/55 | 68.7(9.9) | 139.9(17.9) | 86.1(9.3) | 30.5(5.6) |
| Overall | 800 | 368/432 | 67(9.5) | 136.3(17.5) | 84.6(9.2) | 28.1(4.7) |

**Figure 1:**
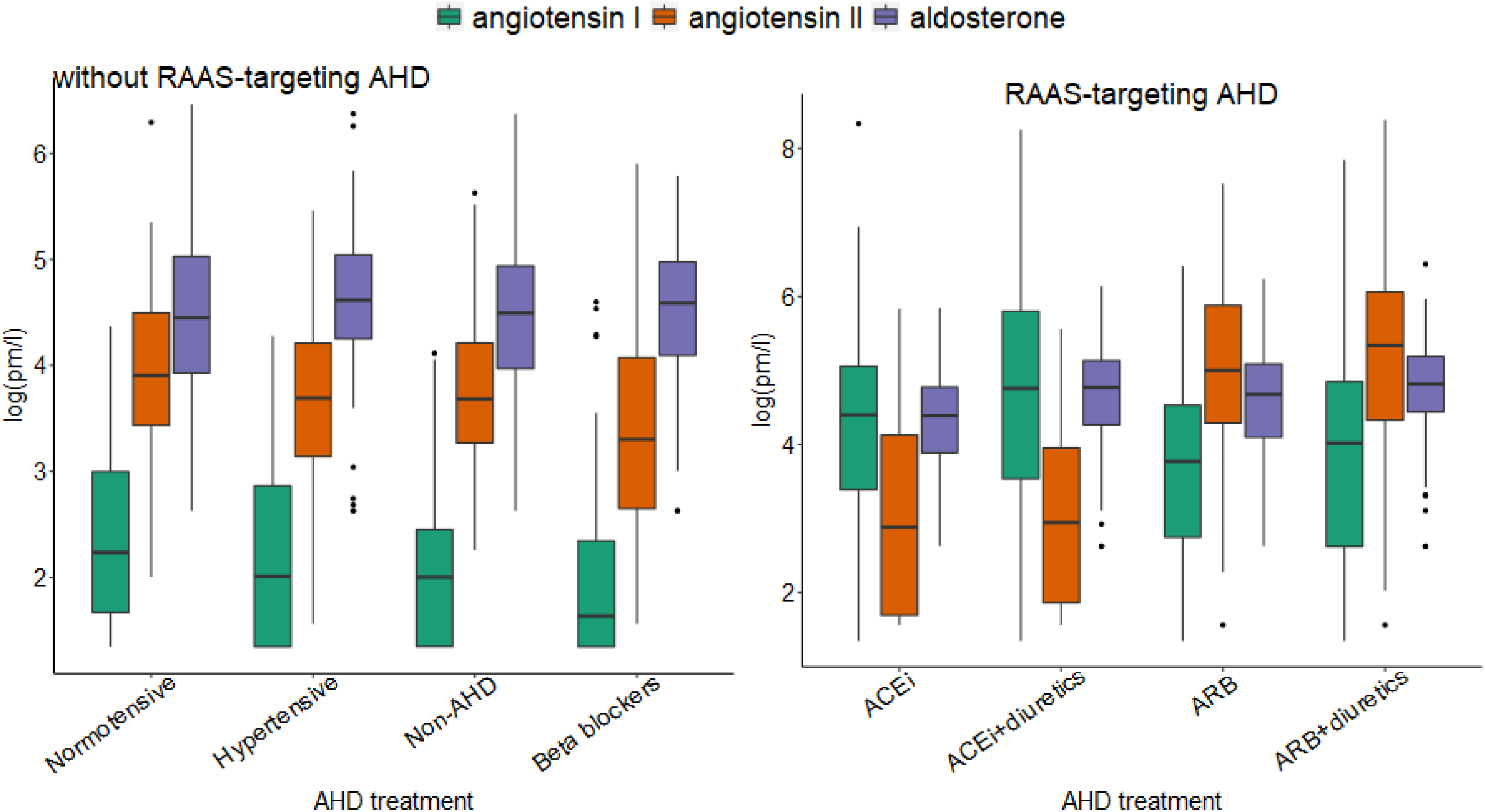
Distribution of angiotensin I, angiotensin II, and aldosterone according to AHD treatment status.

### Cluster analysis

Silhouette index analysis identified the unsupervised k-means clustering method with three clusters as the most appropriate clustering solution as compared to alternative clustering methods (Figure 2a). The k-means cluster analysis of angiotensin I, angiotensin II, and aldosterone identified three well-separated clusters (Figure 2b). The three PCs explained, respectively, 62%, 28%, and 10% of the RAAS biomarkers total variability. Cluster 1, 2, and 3, included 55%, 30% and 15% of the study participants, respectively. The three RAAS biomarkers showed substantially different distributions (one-way ANOVA test P<0.0001) and distinct patterns over the three clusters (Figure 2c): angiotensin I was lowest in cluster 1, intermediate in cluster 2 and largest in cluster 3, with non-overlapping distributions between clusters 1 and 3. Angiotensin II peaked in cluster 2 and showed lowest levels in cluster 3, with nearly non-overlapping distribution between clusters 2 and 3. Aldosterone was relatively less variable across the clusters, yet the difference between the clusters was statistically significant.

**Figure 2:**
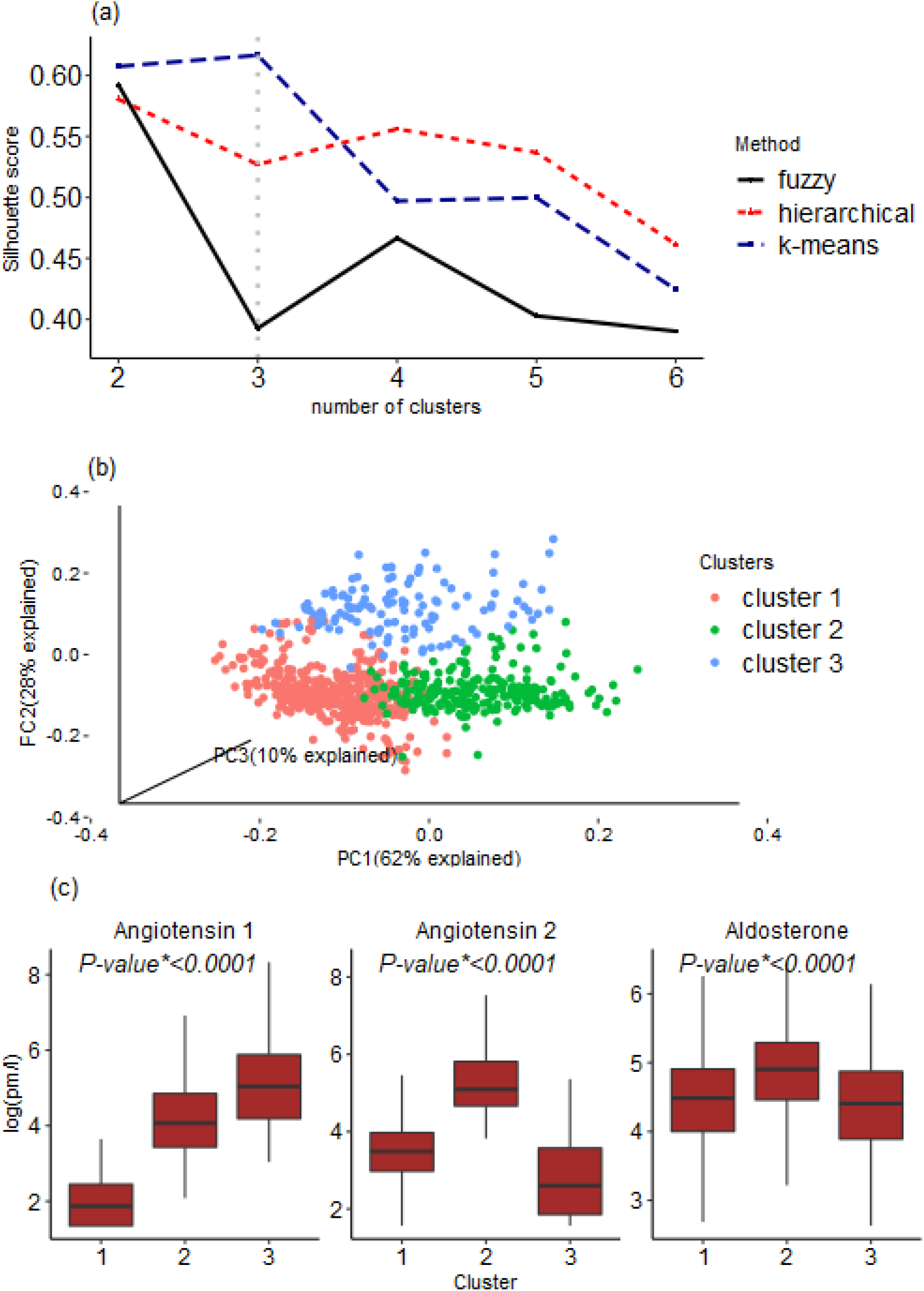
Cluster analysis result. **Panel a**: identification of the most appropriate clustering solution via the Silhouette score metric. **Panel b**: The resulting well-separated clusters; **Panel c**: Distribution of the three RAAS biomarkers across clusters. P-value* from one-way ANOVA test.

### Identification of AHD group by Clusters

We evaluated to what extent the three clusters were able to identify different AHD treatment groups. Figure 3 depicts the study participants stratified by the eight AHD groups and by clusters. Cluster 1 comprised individuals from all groups but with strong preponderance of individuals from the normotensive, hypertensive, non-AHD, and beta blockers groups, that is, cluster 1 seems to represent individuals who are not on RAAS-targeting treatment (ATC=C09). Cluster 2 was enriched for individuals on ARB with or without diuretics. Cluster 3 included only individuals on ACEi with or without diuretics.

**Figure 3:**
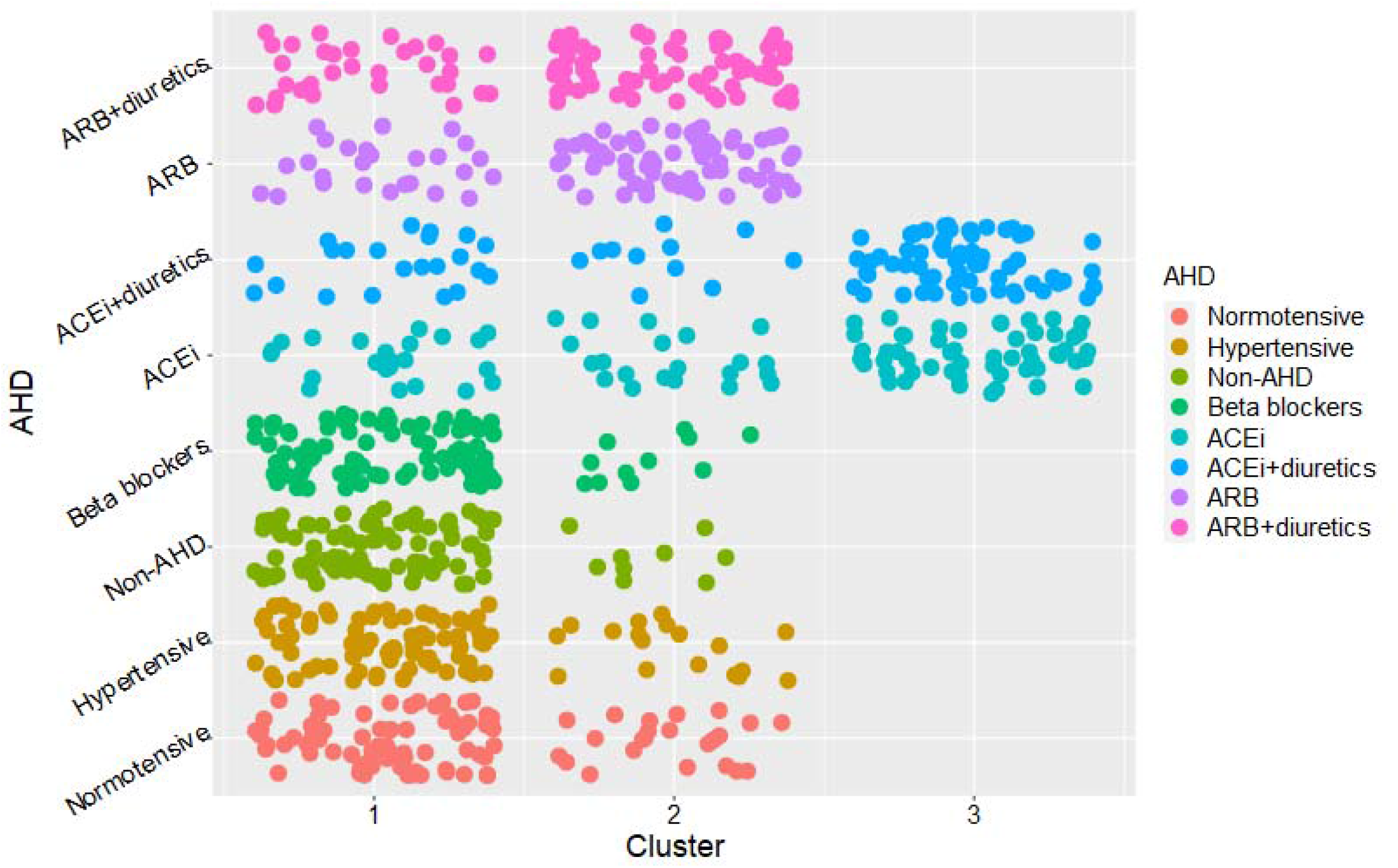
Distribution of study participants across eight AHD treatment status groups against the three identified clusters.

Results from the analysis of the classification properties of the three clusters are shown in Table 2. We observed highest agreement between cluster 3 and ACEi (κ_w_=82%), with 55% sensitivity and 90% specificity, and between cluster 3 and ACEi+diuretics (κ_w_=78%, sensitivity=67%, specificity=92%). When joining ACEi and ACEi+diuretics groups together, cluster 3 showed κ_w_ =82% (95%CI: 79%-86%), 61% (95%CI: 54%-68%) sensitivity, and 100% (95%CI: 99%-100%) specificity. Cluster 2 had best agreement with ARB (κ_w_=74%, sensitivity=73%, specificity=83%) and ARB+diuretics (κ_w_=51%, sensitivity=69%, specificity=76%). When joining ARB and ARB+diuretics groups together, cluster 2 showed 71% (95%CI: 64%-77%) sensitivity and 84% (95%CI: 81%-87%) specificity. Cluster 1 showed a sensitivity between 77% and 91% to identify individuals from the normotensive, hypertensive, non-AHD and beta blocker groups (sensitivity=85% when joining the four groups together). Specificity of cluster 1 to assess that an individual is either normotensive, hypertensive, non-AHD or beta blocker user but not a RAAS AHD user was 74% (95%CI: 69%-78%).

**Table 2:** Weighted kappa, sensitivity and specificity for identification of AHD treatment between RAAS-generated clusters and AHD classification via scanning drug boxes.

| AHD group | Weighted kappa |  |  | Sensitivity |  |  | Specificity |  |  |
| --- | --- | --- | --- | --- | --- | --- | --- | --- | --- |
|  | Cluster1 | Cluster2 | Cluster3 | Cluster1 | Cluster2 | Cluster3 | Cluster1 | Cluster2 | Cluster3 |
| Normotensive | 63(58-68) | 46(39-52) | 12(4-19) | 77(68-85) | 23(15-32) | 0(0-4) | 48(44-51) | 70(66-73) | 83(80-85) |
| Hypertensive | 64(59-69) | 44(37-50) | 14(6-21) | 82(73-89) | 18(11-27) | 0(0-4) | 48(45-52) | 69(65-72) | 83(80-85) |
| Non-AHD | 60(56-66) | 41(34-48) | 18(10-25) | 91(84-96) | 9(4-16) | 0(0-4) | 50(46-53) | 68(64-71) | 83(80-85) |
| Beta blockers | 45(39-52) | 22(17-28) | 6(1-12) | 89(81-94) | 11(6-19) | 0(0-4) | 49(46-53) | 49(46-53) | 0(0-4) |
| ACEi | 12(4-20) | 45(39-52) | 82(78-85) | 24(16-34) | 21(14-31) | 55(44-65) | 40(36-44) | 69(66-73) | 90(88-93) |
| ACEi+diuretics | 25(19-31) | 22(17-28) | 78(75-82) | 22(14-31) | 11(6-19) | 67(57-76) | 40(36-43) | 68(64-71) | 92(90-94) |
| ARB | 23(18-29) | 74(69-80) | 45(39-49) | 27(18-36) | 74(64-82) | 0(0-4) | 40(37-44) | 77(73-80) | 83(80-85) |
| ARB+diuretics | 21(15-27) | 51(46-55) | 44(38-51) | 31(23-41) | 69(59-77) | 0(0-4) | 41(37-45) | 76(73-79) | 83(80-85) |
| Non-RAAS* | 23(16-31) | 60(53-65) | 16(8-24) | 85(81-88) | 15(12-19) | 0(0-1) | 74(69-78) | 56(51-61) | 70(65-74) |
| ACEi or ACEi+diur** | 12(4-22) | 45(38-54) | 82(79-86) | 23(17-30) | 16(11-22) | 61(54-68) | 34(30-38) | 66(62-70) | 100(99-100) |
| ARB or ARB+diur*** | 70(64-74) | 41(34-48) | 27(19-34) | 29(23-36) | 71(64-77) | 0(0-2) | 36(32-40) | 84(81-87) | 80(76-83) |

\* participants not on RAAS targeting drugs
\*\*group of participants who received the RAAS targeting ACEi or ACEi + diuretics.
\*\*\*Participants who received the RAAS targeting ARB or ARB+diuretics

### Clinical features of the clusters and association with RAAS biomarkers

The clinical characteristics of each cluster are described in Table 3. Cluster 1 was characterized by higher average SBP and DBP than clusters 2 and 3. Individuals in clusters 2 and 3 displayed more cardiometabolic abnormalities, including elevated fasting blood glucose levels and HbA1c, higher BMI, reduced eGFR levels, and a greater proportion of individuals with DM compared to cluster 1. There were no significant differences in cholesterol, cortisol, sodium or potassium levels among the three clusters.

**Table 3:** Distribution of clinical and laboratory characteristics across the three clusters. Continuous variables are presented as mean (sd); categorical variables are presented as counts and percentages (%).

| Variable | Cluster 1 (n=444) | Cluster 2 (n=235) | Cluster 3 (n=121) | P-value* |
| --- | --- | --- | --- | --- |
|  | Mean(sd) | Mean(sd) | Mean(sd) |  |
| Age (Year) | 66.57(9.1) | 65.71(9.7) | 68.6(10.5) | 0.1826 |
| Sex F: n(%) | 240(54.0) | 129(55.0) | 63(52.0) | 0.8789 |
| BMI (kg/m<sup>2</sup>) | 27.4(4.5) | 29.0(4.9) | 29.3(4.5) | 0.0021 |
| HbA1c(%) | 5.91(0.47) | 6.04(0.63) | 6.04(0.52) | $1.05 \times 10^{-6}$ |
| Glucose (mg/dl) | 98.76(15) | 104.38(23.12) | 102.48(15.69) | $4.42 \times 10^{-5}$ |
| Cholesterol (mg/dl) | 224.19(41.68) | 220.01(44.99) | 221.36(41.83) | 0.3235 |
| eGFR (mL/min) | 78.34(12.47) | 74.51(14.85) | 73.55(15.77) | 0.0070 |
| Sodium (mmol/L) | 141.1(2.19) | 140.73(2.49) | 140.53(2.23) | 0.8505 |
| Potassium (mmol/L) | 4.59(0.38) | 4.58(0.41) | 4.59(0.42) | 0.7828 |
| Cortisol (µg/dl) | 13.53(4.52) | 13.69(4.73) | 13.59(4.89) | 0.0967 |
| SBP (mmHg) | 137.99(18.46) | 132.4(14.63) | 137.54(17.9) | 0.0434 |
| DBP (mmHg) | 85.27(9.4) | 83.56(8.5) | 83.96(9.42) | 0.0019 |
| Diabetes: n(%) | 42(9%) | 51(22%) | 25(21%) | $1.45 \times 10^{-5}$ |

\*From one-way ANOVA for continuous and Chi-square test for categorical variables.

If, after removing both the cluster and the treatment effects, the clinical characteristics considered above were still associated with angiotensin I, angiotensin II and aldosterone levels, that would indicate the presence of variability not captured by the two classifiers (clusters, treatment) and thus possible reasons for imperfect agreement between them. Lasso model fitting results are shown in Table 4. After removing the effect of the clusters and the treatment, angiotensin I was still associated with age, sex, eGFR, DBP, and DM. Angiotensin II was still associated with age, sex, eGFR, and SBP. Aldosterone was associated with sex, BMI, eGFR, and cortisol.

**Table 4:** Clinical predictors of angiotensin I, angiotensin II and aldosterone after accounting for cluster structure and AHD treatment.

| Variables | Angiotensin I |  | Angiotensin II |  | Aldosterone |  |
| --- | --- | --- | --- | --- | --- | --- |
|  | Est(SE) | P-value | Est(SE) | P-value | Est(SE) | P-value |
| Age, year | -0.025(0.005) | $2.17 \times 10^{-6}$ | -0.017(0.005) | 0.0012 | *removed\** | |
| Sex M | 0.416(0.087) | $2.18 \times 10^{-6}$ | 0.308(0.077) | $7.85 \times 10^{-5}$ | -0.222(0.054) | $4.95 \times 10^{-5}$ |
| BMI kg/m<sup>2</sup> | 0.018(0.01) | 0.0666 | 0.014(0.009) | 0.1353 | 0.016(0.006) | 0.0113 |
| HbA1c % | -0.109(0.101) | 0.2839 | *removed\** |  | *removed\** |  |
| Glucose mg/dl | *removed\** |  | 0.001(0.003) | 0.8870 | 0.003(0.002) | 0.1492 |
| Cholesterol, mg/dl | 0.002(0.001) | 0.0743 | *removed\** |  | *removed\** |  |
| eGFR ml/min/1.73m<sup>2</sup> | -0.013(0.004) | 0.0002 | -0.007(0.003) | 0.0399 | -0.006(0.002) | 0.0013 |
| Sodium, mmol/L | -0.034(0.018) | 0.0627 | -0.026(0.016) | 0.1095 | 0.002(0.012) | 0.8324 |
| Potassium, mmol/L | 0.135(0.113) | 0.2356 | 0.119(0.102) | 0.2417 | -0.055(0.072) | 0.4416 |
| Cortisol, µg/dl | *removed\** |  | 0.008(0.008) | 0.3281 | 0.012(0.006) | 0.0443 |
| SBP, mmHg | *removed\** | | -0.015(0.004) | $2.69 \times 10^{-5}$ | *removed\** | |
| DBP, mmHg | -0.029(0.005) | $3.70 \times 10^{-8}$ | -0.007(0.007) | 0.2694 | 0.001(0.003) | 0.7244 |
| DM | 0.303(0.154) | 0.0493 | 0.178(0.13) | 0.1719 | -0.074(0.092) | 0.4215 |

Est: estimate of the coefficient of association; SE: standard error of estimated coefficient
\*removed by the lasso penalization algorithm when a predictor is less influential

## Discussion

We investigated to what extent unsupervised cluster analysis applied to measured RAAS biomarkers may help identify individuals from the general population according to the most likely AHD treatment. Our results show that unsupervised clustering can reliably identify individuals on ACEi monotherapy or in combination with diuretics. To a lower extent, clustering can also identify ARB users, with or without diuretics. This is in line with several studies reporting changes in the biomarkers in response to RAAS targeting treatment [8,25]. Furthermore, normotensive, untreated hypertensive and beta blockers groups were classified in the same cluster 1. This implies that the clustering based on the three biomarkers was able to separate those classified as agents acting on RAAS (ACEi and ARB), from the remaining groups, including beta blockers. Despite beta1-adrenergic receptor blockade suppresses renin release directly acting on juxtaglomerular cells [26], an impact of beta blockers on the analyzed biomarkers was not evident in our study.

Cluster 3, representing ACEi users, showed nearly perfect specificity, meaning that individuals who are not on ACEi would be very unlikely to be falsely classified as ACEi user. On the other hand, cluster 3 exhibited limited sensitivity, with nearly four out of ten ACEi users being missed by this classifier. Cluster 2, mainly representing ARB users, showed higher sensitivity, with seven out of ten ARB users that would be correctly identified by this classifier, but imperfect specificity, allowing some non-ARB user to enter this group.

Clusters 2 and 3, encompassing ARB and ACEi users, displayed a higher rate of cardiometabolic abnormalities, including DM and elevated fasting blood glucose levels and HbA1c, higher BMI, and reduced eGFR levels. This is consistent with current clinical protocols, which prioritize assignment of ACEi and ARB for individuals with metabolic syndrome like diabetic hypertensive [27] and kidney disease [28] patients. The identified clusters were not different in terms of cortisol, potassium and sodium levels. The absence of an association with potassium, whose level can be depleted by thiazide diuretics reflects the inability of cluster analysis to discriminate those on diuretic treatment among those taking ACEi or ARB.

The penalized regression analysis of residual variability after removing the effect of the identified clusters and treatment groups, highlighted residual strong associations with age and sex with all three RAAS biomarkers. Also higher eGFR, which indicates better kidney function, was associated with lower levels of all three RAAS biomarkers. A lower SBP was associated with higher angiotensin II levels, according to expectations since a drop in BP triggers RAAS to increase BP through increased release of angiotensin [29]. The detection of these associations after removal of the treatment effect and of the cluster effect, indicates the presence of additional factors acting on angiotensin I, angiotensin II and aldosterone levels that might explain the imperfect agreement between clusters obtained through unsupervised statistical analysis and objective AHD classification up on participation. In particular, there is a known differential prescription of AHD by sex [30]. However, given we adjusted the analyses for drug groups and groups were sex-matched, it is more likely that the residual association with sex is of purely biological origin.

The main feature of our study was the analysis of RAAS biomarkers typically analysed only in clinical context in a population-based scenario. This was possible thanks a novel quantification method that allows RAAS biomarker quantification in frozen samples, thus allowing use in general population studies conducted outside of controlled clinical settings. On the other hand, limitations should be highlighted. This study relied on cross-sectional measurement which might not be reflective of the actual health status of an individual over an extended period, especially for what concerns BP. Imperfect discrimination by cluster analysis could be explained by heterogeneous counter-regulatory renin release mechanisms and by ACEi/ARB escape phenomenon, the latter dependent on individual drug response [31]. Classification based on barcode scanning of drug boxes provides great precision but our study could not take into account adherence to treatment since drug levels were not measured. This unaccounted variability may have additionally contributed to the imperfect agreement between clusters and drug groups as noncompliance is known to affect sensitivity and specificity of treatment screening [32]. Finally, while we identified a promising unsupervised procedure to identify underlying AHD treatment targeting the RAAS system, and despite submitting the clustering procedure to cross validation, our limited sample size has prevented us an independent replication and calibration of the clustering algorithm in an independent setting is warranted.

In conclusion, our study has demonstrated that mass-spectrometry based assessment of RAAS-biomarkers in previously biobanked samples can provide reliable information to identify RAAS-based AHD treatment in general population studies.

## Data Availability

All data produced in the present study are available upon reasonable request to the authors

## Acknowledgements

The CHRIS study is a collaborative effort between the Eurac Research Institute for Biomedicine and the Healthcare System of the Autonomous Province of Bozen/Bolzano. The investigators thank all study participants from the middle and upper Vinschgau/Val Venosta, the general practitioners, the personnel of the Hospital of Schlanders/Silandro, the field study team and the personnel of the CHRIS Biobank (BRIF code BRIF6107) for their support and collaboration. Extensive acknowledgement is reported at https://translational-medicine.biomedcentral.com/articles/10.1186/s12967-015-0704-9.

## Ethical statement

The CHRIS study was approved by the Ethical Committee of the Healthcare System of the Autonomous Province of Bozen/Bolzano, protocol no. 21/2011 (19 Apr 2011). All participants gave written informed consent.

## CRediT author statement

**Arisido M Woldeyes**: Methodology, Investigation, Software, Formal analysis, Writing - Original Draft, Writing - Review & Editing, Visualization

**Foco Luisa**: Conceptualization, Data Curation, Writing - Review & Editing, Project administration

**Shoemaker Robin**: Writing - Review & Editing

**Melotti Roberto**: Writing - Review & Editing

**Delles Christian**: Conceptualization, Writing - Review & Editing

**Gögele Martin**: Data Curation

**Barolo Stefano:** Conceptualization

**Baron Stephanie:** Conceptualization

**Azizi Michel:** Conceptualization

**Dominiczak Anna:** Conceptualization, Writing - Review & Editing

**Zennaro M Christina**: Conceptualization, Investigation, Writing - Review & Editing

**Pramstaller P Peter:** Resources

**Poglitsch Marko**: Investigation, Resources, Writing - Review & Editing

**Pattaro Cristian**: Conceptualization, Methodology, Writing - Review & Editing, Visualization, supervision, Investigation

### Abbreviations

AHD: Anti-hypertensive drugs
RAAS: Renin-Angiotensin-Aldosterone System
CHRIS: Cooperative Health Research In South Tyrol
ARB: Angiotensin type 1 receptor blockers
ACEi: Angiotensin-converting enzyme inhibitors
BP: blood pressure
DBP: diastolic blood pressure
SBP: systolic blood pressure
eGFR: estimated glomerular filtration rate
CV: coefficient of variation
PC: Principal components
ANOVA: analysis of variance
MSE: mean squared error

