## Supplemental appendix for "Cluster analysis of angiotensin biomarkers to identify antihypertensive drug treatment in population studies"

**Supplementary material**

Summary

### Supplementary Figure 1: Distributions of Angiotensin I, Angiotensin II and Aldosterone in the study sample . The density lines were obtained using the Gaussian smoothing kernel.

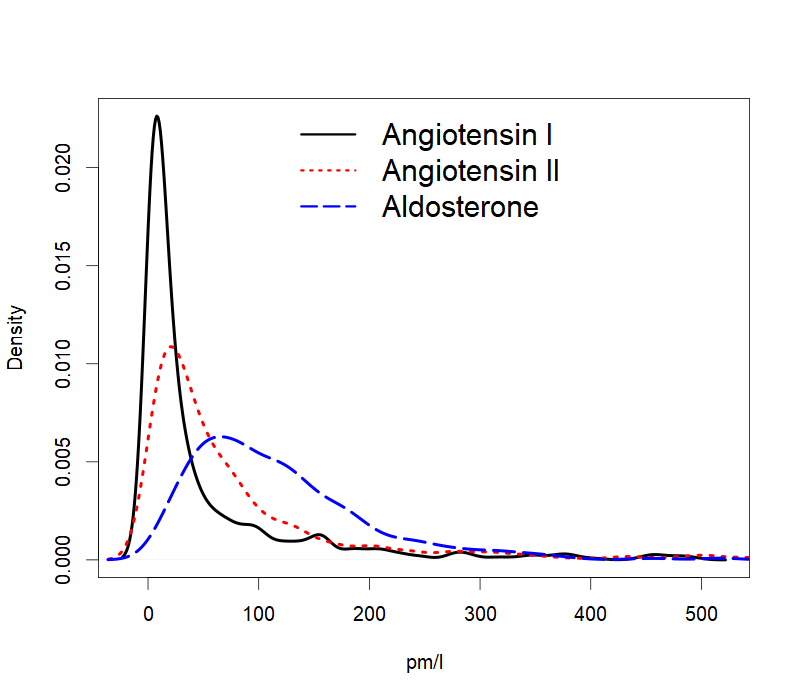

### Supplementary Figure 2: Pairwise correlations between the clinical variables tested for inclusion in the penalized regression model.

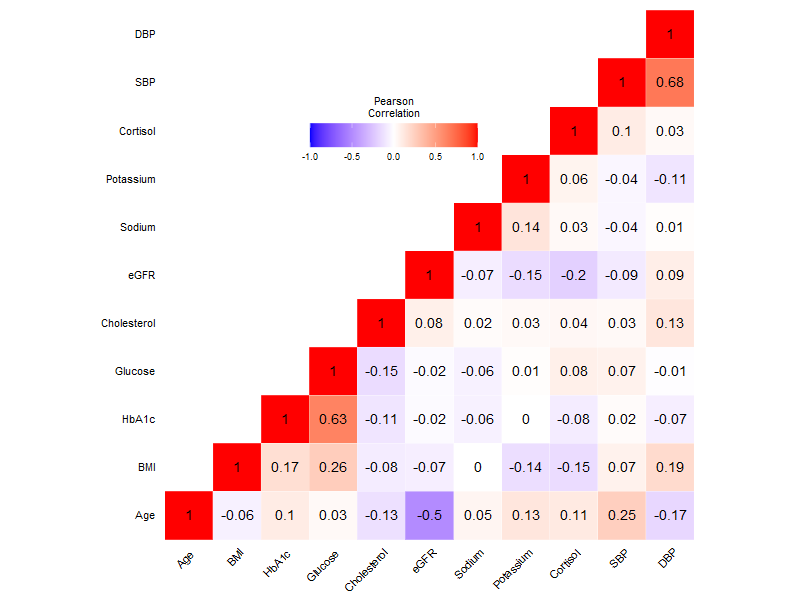

### Supplementary Table 1: Distributions of clinical variables across AHD groups. Reported are means and standard deviations.

| AHD group | Glucose  (mg/dl) | HbA1c  (%) | Cholesterol (mg/dl) | eGFR (mL/min) | Potassium (mmol/L) | Sodium (mmol/L) | Cortisol (mmol/L) |
| --- | --- | --- | --- | --- | --- | --- | --- |
| Normotensive | 96.48(9.8) | 5.86(0.37) | 235.46(40.27) | 78.93(11.13) | 4.67(0.41) | 141.19(2.41) | 14.02(5.42) |
| Hypertensive | 99.19(14.29) | 5.87(0.41) | 236.85(39.68) | 80.68(12.26) | 4.59(0.33) | 141.12(2.09) | 14.01(4.48) |
| Non-AHD | 97.64(13.98) | 5.85(0.46) | 224.15(42.18) | 77.87(13.11) | 4.66(0.38) | 141.25(2.29) | 13.81(4.2) |
| Beta blocker | 99.71(11.62) | 5.93(0.38) | 215.93(44) | 76.8(14.25) | 4.62(0.36) | 140.79(2.08) | 13.41(4.51) |
| ACEi | 102.02(15.99) | 6.08(0.58) | 220.22(44.35) | 74.31(15.75) | 4.73(0.45) | 140.78(2.32) | 13.46(4.89) |
| ACEi + diuretic | 105.15(23.1) | 6.11(0.69) | 215.26(39.4) | 72.68(15.55) | 4.44(0.36) | 140.62(2.31) | 13.75(5.09) |
| ARB | 102.7(28.93) | 6(0.65) | 222.4(39.81) | 76.21(13.8) | 4.6(0.4) | 140.54(1.94) | 12.97(4.14) |
| ARB + diuretic | 104.89(17.4) | 6.08(0.56) | 210.11(45.23) | 74.42(13.31) | 4.41(0.34) | 140.95(2.79) | 13.27(4.26) |

Supplementary Figure 3: Identification of the optimal penalty parameter λ for the lasso regression. **Results are shown from the 8-fold cross-validation (CV). The vertical gray dashed lines represent the penalty parameter that achieved the least MSE.**

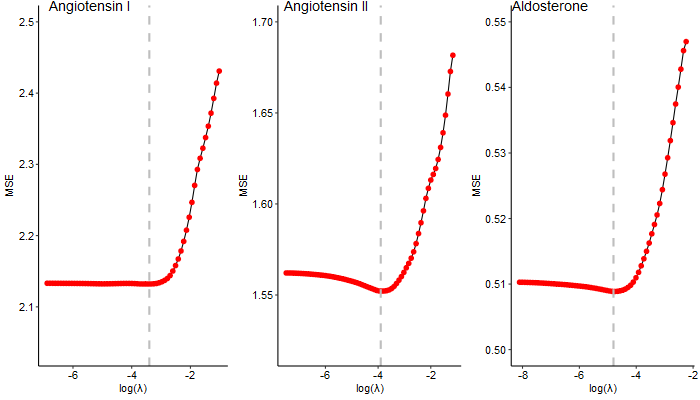
